# Apraxia profiles predict general cognitive deficits in patients with biomarker-verified Alzheimer’s pathology

**DOI:** 10.1101/2024.04.08.24305477

**Authors:** Peter H. Weiss, Claudia C. Schmidt, Michella Barddakan, Elena Jaeger, Nils Richter, Gérard N. Bischof, Kathrin Giehl, Özgür A. Onur, Frank Jessen, Gereon R. Fink, Alexander Drzezga

## Abstract

**Background:** Apraxia represents a core feature of Alzheimer’s disease, a neurodegenerative disorder associated with increased β-amyloid plaques and tau deposition. However, descriptions of apraxic deficits in AD patients are still scarce. Here, we comprehensively investigate apraxia profiles and their impact on general cognitive deficits in patients with biomarker-verified Alzheimer’s pathology.

**Methods:** We characterized the frequency and patterns of apraxic deficits in patients with biomarker-verified Alzheimer’s pathology (n=45) using a battery of standardized apraxia tests. Demographic variables and apraxia scores were related to patients’ general cognitive impairment using hierarchical regression analyses.

**Results:** Apraxic deficits were found in 78% of patients with biomarker-verified Alzheimer’s pathology. AD patients were more impaired in imitating finger gestures (than hand gestures: 76.6% vs. 87.8%, p < 0.001), and imitating complex hand movements (than single hand movements: 76.3% vs. 96.7%, p < 0.001), even when controlling for general cognitive impairment. Apraxia assessments explained about 60% of the variance related to the severity of general cognitive deficits, with deficits in pantomiming object use (beta coefficient: 0.55, p = 0.017) and imitating finger gestures (beta coefficient: −0.51, p < 0.001) being significant predictors of general cognitive impairment.

**Conclusions:** These findings underline the relevance of apraxia in patients with biomarker-verified Alzheimer’s pathology. Data revealed distinct apraxia profiles independent of patients’ general cognitive status and showed that praxis performance, especially apraxic deficits in pantomiming the use of objects and imitating finger gestures, predicts general cognitive functioning in Alzheimer’s disease.

**What is already known on this topic:** Apraxic deficits have been commonly reported in patients with suspected Alzheimer’s disease (AD). Current diagnostic criteria of AD include disease-specific biomarkers (i.e., amyloid and tau) reflecting the neuropathological changes in AD. However, little is known about the prevalence and characteristics of apraxia in patients with biomarker-verified Alzheimer’s pathology.

**What this study adds:** In a well-defined sample of patients with evidence of Alzheimer’s pathology based on positive amyloid and tau biomarkers in cerebrospinal fluid (CSF) or positron emission tomography (PET), apraxic deficits are common, show differential patterns even after controlling for general cognitive deficits, and account for variance in the severity of the patients’ general cognitive impairment.

**How this study might affect research, practice or policy:** This study shows that in patients with biomarker-verified Alzheimer’s pathology apraxic deficits are a relevant symptom that can predict general cognitive performance in mild to moderate disease stages.

Our results warrant further investigation into the neuropathology underlying apraxic deficits in Alzheimer’s disease by examining the relationship between apraxic deficits and regional amyloid and tau deposition in the brain.

## INTRODUCTION

Alzheimer’s disease (AD) is a neurodegenerative disorder that is primarily characterized by a progressive decline in cognitive functions, particularly episodic memory (Ballard et al., 2011). Importantly, current diagnostic criteria for AD indicate the presence of atypical, non-amnestic neuropsychological symptoms of AD (McKhann et al., 2011). Among these symptoms, apraxia stands out as a critical cognitive phenotype that remains under-investigated in AD patients (Lesourd et al., 2013). Apraxia is a disorder affecting cognitive motor functions related to gesture imitation, pantomiming object use and/or actual object use that cannot be (solely) attributed to basic motor deficits (Cubelli, 2017). This disorder constitutes a core feature of AD that likely increases in severity (Parakh et al., 2004) and prevalence (Lesourd et al., 2013) as AD progresses, with the prevalence of apraxic deficits shown to increase from approximately 30% in mild cases of AD to a staggering 90% in severe stages of the disease.

Notably, the prevalence and/or severity of (limb) apraxia have been demonstrated to serve as effective markers in distinguishing AD from frontotemporal dementia (Ahmed et al., 2016; Chandra et al., 2015), as well as from mild cognitive impairment and subcortical vascular dementia (Ozkan et al., 2013). Interestingly, already in the early stages of AD, apraxia supports the differential diagnosis of AD from other subtypes of dementia, since AD patients present with distinctive clinical patterns of apraxic deficits (Johnen et al., 2015a; 2015b; 2018). In particular, patients with AD exhibited pronounced deficits in imitating hand and finger gestures, whereas patients with frontotemporal dementia showed specific impairments in the imitation of bucco-facial gestures (Johnen et al., 2015a). Notably, this specific apraxia profile in AD patients (i.e., more severe deficits in limb apraxia compared with bucco-facial apraxia) also showed an association with cerebrospinal fluid (CSF)-based biomarkers indicative of (additional) AD pathology in patients with frontotemporal dementia (Pawlowski et al., 2018).

Despite the documented importance of apraxic deficits in AD, systematic investigations into the frequency and profiles of apraxic deficits in AD remain sparse, particularly in patients with biomarker-verified Alzheimer’s pathology. According to the current diagnostic guidelines of the National Institute on Aging and the Alzheimer’s Association (NIA-AA; McKhann et al., 2011), the identification of Alzheimer’s disease hinges on the detection of specific neuropathological markers, namely amyloid-β (A+) and tau (T+) that are detected by analysis of CSF or Positron Emission Tomography (PET; Dubois et al., 2014). Importantly, this biomarker-based definition of AD is considered to be crucial in distinguishing AD from other neurodegenerative disorders leading to dementia (Jack et al., 2018).

The objective of the present study was to comprehensively characterize apraxic deficits in a well-defined sample of patients with Alzheimer’s pathology, as evidenced by CSF or PET derived biomarkers (i.e., amyloid-β (A+) and tau (T+)). In particular, we investigated the frequency and severity of apraxic deficits as well as their relationship to the general cognitive status of the patients. Moreover, we elucidated the presence of differential patterns of praxis deficits (i.e., apraxia profiles) in patients with Alzheimer’s pathology, while controlling for their general cognitive deficits. Specifically, our study investigated whether patients with biomarker-verified Alzheimer’s pathology exhibit greater deficits in imitating hand or finger gestures, and whether their impairments are more pronounced in imitating complex versus simple gestures. Finally, by examining which apraxic deficits predict general cognitive impairments in patients, this study seeks to provide insights into the potential predictive value of praxis performance for cognitive functioning in patients with biomarker-verified Alzheimer’s pathology.

## MATERIAL AND METHODS

### Patient sample

A total of 78 patients with clinically suspected AD were recruited from the Center for Memory Disorders (ZfG; ‘Zentrum für Gedächtnissstörungen’) run by the Neurology and Psychiatry Departments of the University of Cologne. Patients had to be at least 50 years old and had to fulfill the NIA-AA-criteria for the diagnosis of Alzheimer’s clinical syndrome (McKhann et al., 2011). Patients were excluded (i) if they fulfilled the criteria for other forms of dementia than of the Alzheimer’s disease type or (ii) if they suffered from other disorders potentially responsible for cognitive decline or motor deficits (e.g., cerebrovascular disorders, Parkinson’s disease, Multiple Sclerosis) or (iii) lacked the ability to provide informed consent.

Consequently, the current study included 45 patients (20 females, 44%) with biomarker verified Alzheimer’s pathology. The latter was based on cerebrospinal fluid (CSF) analysis as well as Positron Emission Tomography (PET) scans revealing abnormal amyloid-β (A+) and tau protein (T+) levels.

The patient group had a mean age of 70.0 years (standard deviation [SD] = 9.9, range 50–85) and a mean education level of 14.6 years (SD = 3.6, range 8–24). The patients’ overall cognitive status was assessed using the Mini-Mental State Examination (MMSE), a test that evaluates impairments in general cognitive functions including orientation, attention, working memory, language, and delayed recall (Folstein et al., 1975). Adhering to the classification guidelines proposed by Folstein et al. (2001), the studied patients showed mild (N = 21, 47%) to moderate (N = 14, 31%) general cognitive decline as indicated by an MMSE score of 20-26 and 10-19 points, respectively. Ten patients (22%) showed no signs of general cognitive impairment according to an MMSE score above 26 points. Notably, none of the current patients was classified as having severe cognitive decline (MMSE < 10).

The study was approved by the local ethics committee. Informed consent was obtained from all patients.

### Apraxia assessment

The assessment of apraxia was conducted using six different apraxia tests that assess various praxis functions and deficits thereof, namely, the Cologne Apraxia Screening (Kölner Apraxie Screening, KAS; Dovern et al., 2012), the Dementia Apraxia Test (DATE, Johnen et al., 2016), Goldenberg’s tests of imitating hand positions and finger configurations (Goldenberg et al., 1996), De Renzi’s imitation test (De Renzi et al., 1980), and De Renzi’s test for actual object use (De Renzi et al., 1968).

The KAS is a standardized, validated diagnostic tool designed for stroke patients (Weiss et al. 2013), which has also been effectively used to diagnose apraxia in mild dementia cases (Johnen, et al., 2018). It comprises two subtests for assessing pantomimes of object use and two subtests for assessing imitation. In the pantomime subtests, patients are instructed to perform (i) five pantomimes of object use involving bucco-facial aspects additionally to arm/hand movements (e.g., pantomiming the use of a toothbrush), and (ii) five pantomimes of object use involving (solely) arm/hand movements (e.g., pantomiming the use of scissors). In the imitation subtests, patients are instructed to imitate ‘as if seen in the mirror’ (i) five bucco-facial gestures (e.g., sticking out one’s tongue) and (ii) five arm/hand gestures (e.g., making the stop sign with one’s hand). The maximum total KAS score is 80 points (20 points for each subtest), with a score of 76 points or less indicating apraxia (Weiss et al., 2013). For additional information on the quality standards and testing procedure of the KAS, please refer to Dovern et al. (2012) and Schmidt et al. (2022).

The Dementia Apraxia Test (DATE) is a reliable clinical rating tool for evaluating praxis impairments in neurodegenerative diseases (Johnen et al., 2016). The DATE is structured into five subsets, with two assessing limb-related apraxia and three covering bucco-facial related apraxia. The limb-related subset includes (i) eight items involving the imitation of hand and finger postures and (ii) two items of pantomiming object use (single and multiple object use). For bucco-facial apraxia, the subset is comprised of (iii) six items of imitating face postures, (iv) two items of producing emblematic bucco-facial gestures upon verbal command (e.g., ‘show me how you whistle’) and (v) two items aimed at evaluating apraxia of speech by repeating multi-syllabic pseudo-words. The maximum total DATE score is 60 points (30 points for each subdomain), with scores below 45 points indicating the presence of apraxia. For further details on the DATE’s testing procedure, please refer to Johnen et al. (2015a).

Goldenberg’s imitation tests involve reproducing ten hand positions and ten finger configurations demonstrated by an examiner. Each test has a maximum score of 20, with scores below 17 for hand positions and below 18 for finger configurations indicative of apraxic imitation deficits (Goldenberg, 1996). Additionally, De Renzi’s imitation test comprises two subtests evaluating varying levels of sequence complexity. The simple subtest assesses the imitation of single hand positions and finger configurations across 12 items, while the complex subtest comprises the imitation of complex sequences of hand positions and finger configurations, also across 12 items (De Renzi, 1980). The test’s maximum score is 72 points (36 points for each subtest), with a score below 53 indicating apraxia. In addition, the De Renzi test for actual object use was implemented to evaluate the ability to use five single tools (e.g., hammer) and two tool-object pairs (e.g., match and candle), with scores below 30 (out of a total of 32 points) indicating tool-use apraxia.

To ensure comparability across the different apraxia tests, all scores were normalized to percentage values, where a score of 100% represents the maximum achievable score on a given test.

### Statistical analysis

Statistical analyses were conducted using the IBM SPSS Statistics (Statistical Package for the Social Sciences, version 25). An overall apraxia severity index was computed for each patient based on the number of impaired apraxia assessments (i.e., the KAS, the DATE, Goldenberg’s tests of imitation, De Renzi’s tests of imitation and actual object use). The apraxia severity index reflected the extent of apraxia impairment, categorizing patients into four distinct groups: patients exhibiting no apraxia (0 tests impaired), mild apraxia (1-2 tests impaired), moderate apraxia (3-4 tests impaired), and severe apraxia (5-6 tests impaired).

Pearson correlation coefficients were calculated to explore the relationship between overall apraxia severity and demographic factors (age and years of education) as well as general cognitive deficits (as indexed by the MMSE score) in the current sample of patients with biomarker verified AD pathology. To elucidate putative dissociations between apraxic impairments and cognitive decline in patients with AD pathology, a chi-square analysis was executed to assess the association between the presence of apraxia (defined by at least one impaired apraxia test) and the presence of cognitive decline (defined by a MMSE score below 27 points).

Building on the existing literature highlighting more pronounced deficits in imitating limb-related gestures compared to facial gestures in patients with AD (Johnen et al., 2015a), the following analyses aimed to further elucidate nuanced impairments in the imitation of the hand and finger gestures. We conducted an Analysis of Covariance (ANCOVA) to investigate putative differential deficits in the imitation of finger versus hand gestures in the current patients with AD pathology. Specifically, the analysis evaluated the overall performance in Goldenberg’s imitation tests (dependent variable), incorporating the within-subject factor of effector (hand versus finger) while controlling for general cognitive deficits (as operationalized by the MMSE score). An additional ANCOVA was conducted to further investigate putative differential deficits in the imitation of complex versus single finger and hand gestures in patients with AD pathology. In particular, the analysis evaluated the overall performance in De Renzi’s imitation test (dependent variable), incorporating the within-subject factors of effector (hand versus finger) and complexity (single versus complex), while also adjusting for general cognitive deficits (as operationalized by the MMSE score). Note that for the second model, one patient was omitted from the analysis due to missing data related to the De Renzi test of imitation.

To investigate whether variance in general cognitive deficits in patients with AD pathology can be explained by demographic factors (i.e., age and education) or by the performance in different subtests of the apraxia assessment, we performed a hierarchical linear regression analysis. In this analysis, two models were defined with the MMSE total score as the dependent variable. The first-level model included age and years of education as predictive variables. In the second-level model, the scores (in %) in the two KAS subtests of pantomiming object use and imitating gestures, the two DATE subtests for limb and bucco-facial apraxia, the two De Renzi subtests of imitating single and complex gestures, and the two Goldenberg tests of imitating hand positions and finger configurations were simultaneously added to the regression model. Note that De Renzi’s object use test was excluded from this analysis as it was identified as the least sensitive in detecting apraxia (see results section ‘*Frequency and severity of apraxic deficits*’). In addition, a complementary hierarchical linear regression analysis was conducted to investigate whether variance in general cognitive deficits in patients with AD pathology can be explained by demographic factors (i.e., age and education) *or* specific apraxic deficits. In particular, the second level of this model included the difference scores between hand and finger imitation in Goldenberg’s imitation tests (computed as: hand – finger imitation) and the interaction scores of the differential impairment in imitating single versus complex hand and finger gestures in the De Renzi imitation test [computed as (hand _single_ – hand _complex_) - (finger _single_ – finger _complex_)].

The fit of the hierarchical linear regression models was estimated by calculating the Akaike Information Criterion (AIC), a measure that assesses goodness of fit while correcting for model complexity by penalizing the inclusion of an increasing number of predictors (Akaike, 1975). This approach allowed a comparison of the first and second levels of each of the two regression models, verifying that any increase in predictive power in the second-level model was not merely attributable to an increase in the number of predictors. Reported significant effects were followed by pairwise comparisons across the factors’ levels using paired samples t-tests with post-hoc Bonferroni corrections for multiple comparisons and a significance level of *p* < 0.05.

## RESULTS

### Frequency and severity of apraxic deficits

In the current cohort of 45 patients with Alzheimer’s pathology, a substantial portion of patients (78%, N = 35) exhibited specific apraxic deficits related to limb and/or bucco-facial apraxia (i.e., were impaired in at least one of the administered apraxia assessments; see Table 1 for an overview of the patients’ performance on the different apraxia assessments). Among these patients with apraxia, 57% (N = 20) exhibited mild apraxia (i.e., impaired in up to 3 apraxia tests), while the remaining 43% (N = 15) suffered from moderate to severe apraxia (impaired in four to six apraxia tests). Notably, the KAS and the DATE emerged as the most sensitive in identifying apraxia within our sample of patients with biomarker-verified AD pathology. Of the 35 patients diagnosed with limb and/or bucco-facial apraxia, the KAS and the DATE detected 31 (88.6%) and 21 patients (60%) with apraxia, respectively. In contrast, the De Renzi test for actual object use showed the least sensitivity, detecting apraxic deficits in only four patients (out of 35 patients, 11.4%).

**Table 1:** Praxis assessments in patients with AD pathology.

| **Apraxia assessment** | **Patients with AD pathology (n = 45)** | **Cut-off scores** | **Number of patients impaired** |
| --- | --- | --- | --- |
| De Renzi test of actual object use | 96.7 ± 10.8<br>(31.3 – 100) | < 94% | 4 |
| De Renzi imitation test | 78.3 ± 16.9<br>(8.3 – 100)<sup>#</sup> | ≤ 72.2% | 14 |
| Goldenberg hand imitation test | 87.8 ± 15.7<br>(20 – 100) | < 90% | 14 |
| Goldenberg finger imitation test | 76.6 ± 24.7<br>(0 – 100) | < 85% | 19 |
| DATE (total score) | 71.9 ± 19.0<br>(15.8 – 100)<sup>#</sup> | ≤ 75% | 21 |
| KAS (total score) | 86.2 ± 16.1<br>(25 – 100) | ≤ 95% | 31 |

Summary of the overall performance of patients with Alzheimer’s disease (AD) pathology across the six apraxia assessments.
The depicted descriptive statistics include the mean, standard deviation, and range. All scores were transformed into percentages (i.e., a score of 100% represents the maximum achievable score on a given test) to ensure comparability across the different apraxia tests. <sup>#</sup> n = 44
DATE: Dementia Apraxia Test, KAS: Kölner (Cologne) Apraxia Screening.

Note that the KAS was originally developed to assess apraxic deficits in stroke patients. Therefore, we here relate the KAS results to the scores of the DATE, which is a dedicated test to diagnose apraxia in different forms of dementia. Specifically, a significant positive correlation (r = 0.794, *p* < 0.001) was detected between apraxia severity as indexed by the KAS total score (in %) and the one indexed by the DATE total score (in %) across the whole sample of patients with AD pathology (N = 44; note that a patient did not perform the DATE). Furthermore, a chi-square analysis revealed a significant association between the classification of apraxia based on the KAS (scores ≤ 95%) and DATE (scores ≤ 75%) tests (*χ^2^* = 5.69, *p* = 0.024) in patients with AD pathology. In particular, in the whole sample of patients with AD pathology, 41% (N = 18) were classified as apraxic according to both the DATE and KAS criteria. About 86% of the 21 patients deemed apraxic according to the DATE also met the criteria for apraxia on the KAS, whereas 60% of the 30 apraxic patients according to the KAS were also classified as apraxic according to the DATE. The analysis also highlighted that 26.6% (N = 12) of all patients showed apraxia on the KAS but not on the DATE, while only 6.6% (N = 3) of all patients exhibited the reverse pattern. In addition, 24.4% (N = 11) of the whole sample were classified as non-apraxic according to both the DATE and KAS assessments.

### Relationship between apraxia and general cognitive impairment

The overall apraxia severity (indexed by the number of impaired tests) significantly correlated with the severity of general cognitive deficits (indexed by the MMSE score) in the current patients with biomarker-verified AD pathology (N = 45). Specifically, more severe cognitive deficits (reflected by lower MMSE scores) were associated with a higher number of impaired apraxia tests (*r* = −0.534, *p* < 0.001). Note that apraxia severity did not correlate with age (*r* = 0.093, *p* = 0.543) or years of education (*r* = 0.018, *p* = 0.905) in the current patient sample.

Importantly, a chi-square analysis revealed a potential dissociation between the presence of apraxia (defined by at least a single impaired apraxia test) and the presence of cognitive decline (defined by a MMSE score below 27 points) within the current patient sample (*χ^2^* = 0.45, *p* = 0.668). While 62.2% of the patients (N = 28) exhibited both apraxia and cognitive decline, cases of apraxia without cognitive decline, as well as cognitive decline without apraxia, were each observed in approximately 15.6% of the cohort (N = 7). Furthermore, a small fraction of the current patients with biomarker-verified AD pathology (6.6%, N = 3) displayed neither cognitive decline nor apraxia (see Table 2).

**Table 2:** Relationship between apraxia and cognitive decline.

|  |  | Cognitive decline<br>(*MMSE* < 27 points) |  |
| --- | --- | --- | --- |
|  |  | No | Yes |
| Apraxia<br>(*impairment in at least one apraxia test*) | No | 3 | 7 |
|  | Yes | 7 | 28 |

Illustration of the overlap in the frequencies of patients with biomarker-verified AD pathology, in whom apraxia (defined as an impairment in at least one apraxia test) and/or cognitive decline (defined by a MMSE score below 27 points) is either present or absent.
A chi-square analysis revealed a potential dissociation between the presence of apraxia and the presence of cognitive impairment in the current patient sample ( $\chi^2 = 0.45$ , $p = 0.668$ ).
While 62.2% of the patients ( $N = 28$ ) exhibited both apraxia and cognitive decline, cases of apraxia without cognitive decline, as well as cognitive decline without apraxia, were each observed in approximately 15.5% of the cohort ( $N = 7$ ). Furthermore, a small fraction of the sample (6.6%, $N = 3$ ) displayed neither cognitive decline nor apraxia.
MMSE: Mini Mental State Examination.

### Specific apraxia profiles (controlled for general cognitive deficits)

The ANCOVA analysis conducted to discern putative differential deficits in the imitation of finger versus hand gestures among patients with biomarker-verified AD pathology (N = 45) yielded a significant main effect of Effector (see Figure 1A) revealing a better performance in the imitation of hand gestures compared to finger gestures (87.8% vs. 76.6%, *p* < 0.001) in patients with AD pathology, even after adjusting for general cognitive deficits (as operationalized by the MMSE score).

**Figure 1:**
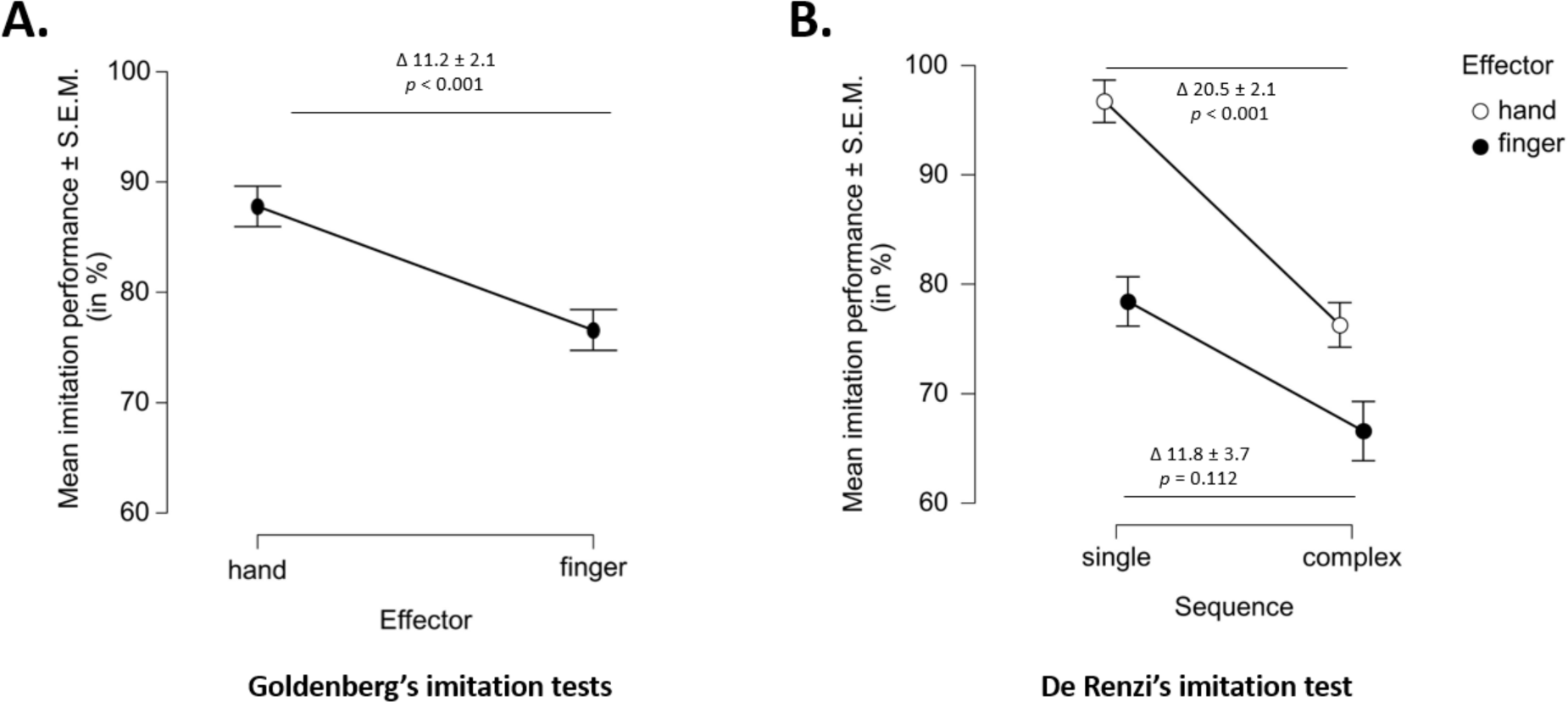
Specific apraxia profiles of the patients with biomarker-verified Alzheimer’s disease pathology (controlled for general cognitive deficits) **Figure 1A** illustrates the higher performance (in %) in imitating hand gestures compared to finger gestures in Goldenberg’s imitation tests among the patients with AD pathology (N = 45). This outcome remained significant even after controlling for general cognitive deficits (as operationalized by the MMSE score). **Figure 1B** illustrates the better performance the patients with AD pathology (N = 44) when imitating hand gestures compared to finger gestures and when imitating single gestures compared to complex gestures in the De Renzi imitation test. In addition, patients exhibited differential deficits when imitating complex hand gestures compared to single hand gestures, whereas the difference in performance between single and complex finger gestures did not reach significance. These outcomes also remained significant even after controlling for general cognitive deficits. SEM: standard error of mean.

Similarly, the ANCOVA investigating putative differential deficits in the imitation of complex versus single (finger and hand) gestures in patients with biomarker-verified AD pathology (N = 44) yielded a significant main effect of Effector, indicating a superior performance in the imitation of hand gestures compared to finger gestures (86.5% vs. 72.5%, *p* = 0.047) as well as a significant main effect of Complexity, indicating a better performance in the imitation of single gestures compared to complex gestures (87.6% vs. 71.4%, *p* < 0.001), even after controlling for general cognitive deficits (as operationalized by the MMSE score) in patients with AD pathology (see Figure 1B). Notably, a significant interaction effect between Effector and Complexity (*p* = 0.02) was also observed after controlling for general cognitive deficits. The interaction term revealed differentially lower scores in imitating complex hand gestures compared to single hand gestures (score difference: 20.5, SD = 2.1; *p* < 0.001), whereas the performance difference between imitating single and complex finger gestures did not reach statistical significance (score difference: 11.8, SD = 3.7, *p* < 0.112).

### Predicting general cognitive impairment by apraxic deficits

The hierarchical regression analysis investigating the extent to which variance in general cognitive deficits could be accounted for by demographic factors (age, education) or by apraxia assessments revealed the importance of the latter. The first-level model, incorporating only age and education did not significantly predict the severity of cognitive deficits in the current patients with AD pathology [adjusted R^2^ = 0.03, F(2,41) = 1.69, *p* = 0.197]. However, the inclusion of the scores obtained in the subtests of the apraxia assessments (two KAS subtests, two DATE subtests, two De Renzi imitation subtests, and two Goldenberg imitation subtests), alongside age and education in the second-level model, significantly improved the model’s predictive capability [adjusted R^2^ = 0.59, F(10,33) = 7.21, *p* < 0.001; R^2^ change: F(8,33) = 8.01, *p* < 0.001]. Among the ten predictors included in the second-level model, only the performance in the KAS subtest assessing pantomiming of object use emerged as an independent predictor of the severity of cognitive deficits (β = 0.55, t = 2.52, *p* = 0.017). Thus, worse performance in the KAS subtest of pantomiming the use of objects predicted more pronounced general cognitive impairment (i.e., lower MMSE scores) in patients with bio-marked verified AD pathology.

Notably, the comparison of the models’ fits using the AIC revealed that the first-level model, limited to only two predictors, had an AIC of 149.08, while the second-level model, featuring ten predictors, achieved a lower AIC of 117.62, and hence, a superior model fit. This observation underscores that the increased predictive power of the second-level model was not simply due to the benefit of added variables *per se*, but was specific to the apraxia subtests additionally included in the model.

The complementary hierarchical regression analysis investigating the extent to which variance in general cognitive deficits in patients with AD pathology could be accounted for by demographic factors or specific apraxic deficits also produced significant results. The first-level model, incorporating only age and education did not significantly predict cognitive deficit severity [adjusted *R*^2^ = 0.03, *F*_(2,41)_ = 1.69, *p* = 0.197]. However, the inclusion of the parameters reflecting the observed specific apraxic deficits in the patients with biomarker-verified AD pathology (the difference scores between hand and finger imitation in Goldenberg’s imitation tests and the interaction scores of the differential impairment in imitating single versus complex hand and finger gestures in the De Renzi imitation test) alongside age and education in the second-level model significantly improved the model’s predictive capability [adjusted *R*^2^ = 0.36, *F*_(4,39)_ = 6.92, *p* < 0.001; *R*^2^ change: *F*_(2,39)_ = 11.30, *p* < 0.001]. Among the four predictors included in the second-level model, only the difference score in the Goldenberg imitation tests emerged as an independent predictor of the severity of cognitive deficits (β = −0.51, *t* = −4.05, *p* < 0.001). Thus, patients with a worse performance in imitating finger gestures compared to hand gestures (i.e., a larger positive difference score) exhibited more severe general cognitive deficits (i.e., lower MMSE scores).

Notably, the comparison of the models’ fits using the AIC revealed that the first-level model, limited to only two predictors, had an AIC of 149.08, while the second-level model, featuring four predictors, achieved a lower AIC of 132.96, and hence, a superior model fit. This observation underscores that the increased predictive power of the second-level model was not simply due to the benefit of added variables *per se*, but was specific to the apraxic deficits.

## DISCUSSION

Our study underscores the high prevalence of apraxia in patients with biomarker-verified AD pathology (A+T+), revealing that a substantial portion (78%; 35 out of 45 patients) of the studied patient sample exhibited apraxic deficits in at least one apraxia test. Moreover, correlation analysis revealed an association between the severity of apraxia impairments and general cognitive deficits (as measured by the MMSE) in patients with AD pathology. Interestingly, the presence of apraxia and cognitive decline was also shown to dissociate in patients with AD pathology. Employing diverse apraxia assessments, we disclosed specific apraxia profiles in patients with AD pathology: AD patients exhibited more pronounced impairments in imitating finger configurations compared to hand gestures as well as in imitating complex compared to simple hand gestures. Importantly, our results highlight that apraxia assessments, rather than demographic factors, significantly accounted for the variability in general cognitive deficit severity within the studied cohort of patients with AD pathology. Specifically, the KAS subtest of pantomiming object use along with differential apraxic deficits in imitating finger gestures, were identified as independent predictors of the severity of cognitive impairments.

Our findings contribute to the understanding of apraxic impairments as potential markers of cognitive decline in AD by demonstrating a significant positive correlation between the severity of apraxic impairments and general cognitive deficits in patients with AD pathology. This observation aligns with previous research indicating an increase in the prevalence of apraxia with increasing dementia severity, as assessed by the MMSE (Parakh et al., 2004). Note that these authors did not report the diagnostic criteria applied in their AD patients. Importantly, the observation that apraxia and cognitive decline also dissociated in some of the current patients with AD pathology further suggests that the manifestation and severity of apraxic symptoms may not consistently relate to the cognitive decline trajectory in all AD patients. Notably, the observation that 7 patients (15%; out of 45) exhibited apraxia without concurrent cognitive decline emphasizes that while apraxia is often associated with cognitive decline, it can manifest independently of it, potentially contributing to the heterogeneity of AD and its atypical, non-amnestic neuropsychological symptoms.

The identification of the KAS subdomain of pantomiming the use of objects as the sole significant predictor, among various assessed praxis domains, of general cognitive deficits in patients with AD pathology, highlights a more nuanced association of cognitive decline with apraxic deficits in AD, specifically with deficits of pantomiming object use. This finding is consistent with previous studies reporting more pronounced pantomime deficits in moderate stages of clinically suspected AD compared to mild stages of the disease (Parakh, 2004; Edwards et al., 1991; Travniczek-Marterer et al., 2009). Note that pantomiming object use is considered a cognitively demanding task that necessitates the retrieval of object-related semantic knowledge (i.e., what an object is used for) and motor schema (i.e., how an object is used) before action execution (Niessen et al., 2014; Roy and Hall, 1992). Accordingly, the predictive value of deficits in pantomiming object use for cognitive decline in AD might potentially arise from its dependence on preserved access and retrieval of conceptual knowledge about object functions and usage (Lesourd et al., 2013). Crucially, both of these cognitive domains are known to be substantially impaired in patients with AD, even in the early stages of the disease (Adlam et al., 2006; Corbett et al., 2012), with several longitudinal studies highlighting semantic knowledge as an early cognitive marker for the conversion from mild cognitive impairment to AD (Vita et al., 2014; Marra et al., 2021).

In addition, the temporal and parietal cortices, which are associated with the storage of conceptual knowledge about object function and use, respectively (Hodges et al., 2000; Kleineberg et al., 2018), are among the regions early affected by neurodegeneration in AD (Whitwell et al., 2010). Crucially, gray matter atrophy in the right middle temporal gyrus and the angular gyrus has been shown to correlate with pantomime performance in patients with early-stage AD (Johnen et al., 2016). Thus, impairments in pantomiming object use not only hold a significant predictive value for cognitive decline in AD, but may also constitute a potential early behavioral marker of AD neuropathology.

## Acknowledgements/ Study funding

This work was funded by the Deutsche Forschungsgemeinschaft (DFG, German Research Foundation) – Project-ID 431549029 – SFB 1451.

GRF gratefully acknowledges support from the Marga and Walter Boll-Stiftung.

Open access publication funded by the Deutsche Forschungsgemeinschaft (DFG, German Research Foundation) – 491111487

## Data availability

Data are available from the corresponding author upon reasonable request.

## Authorship contributions (CRediT author statement)

**Claudia C. Schmidt:** Conceptualization; Formal analysis; Writing – original draft;

**Michella Barddakan:** Writing – review & editing

**Elena Jaeger:** Investigation; Writing – review & editing

**Nils Richter:** Investigation; Writing – review & editing

**Gérard N Bischof:** Writing – review & editing

**Özgür Onur:** Investigation; Writing – review & editing

**Frank Jessen:** Investigation; Writing – review & editing

**Gereon R. Fink:** Resources; Writing – review & editing; Project administration; Funding acquisition

**Alexander Drzezga:** Conceptualization; Writing – review & editing; Supervision; Project administration; Funding acquisition

**Peter H. Weiss:** Conceptualization; Resources; Writing – review & editing; Supervision; Project administration; Funding acquisition

